# Population perspective comparing COVID-19 to all and common causes of death in seven European countries

**DOI:** 10.1101/2020.08.07.20170225

**Authors:** Bayanne Olabi, Jayshree Bagaria, Sunil S. Bhopal, Gwenetta D. Curry, Nazmy Villarroel, Raj Bhopal

**Affiliations:** Department of Dermatology, Lauriston Building, Lauriston Place, Edinburgh, EH3 9EN; Independent Consultant in Global Public Health; Population Health Sciences Institute, Newcastle upon Tyne, NE1 7RU; Usher Institute, University of Edinburgh, EH3 9AG; University of Alabama, Tuscaloosa, AL 35487; University of Sheffield, S10 2TG

## Abstract

**Background:** Mortality statistics on the COVID-19 pandemic have led to widespread concern and fear. To contextualise these data, we compared mortality related to COVID-19 with all and common causes of death, stratifying by age and sex. We also calculated deaths as a proportion of the population by age and sex.

**Methods:** COVID-19 related mortality and population statistics from seven European countries were extracted: England and Wales, Italy, Germany, Spain, France, Portugal and Netherlands. Available data spanned 14-16 weeks since the first recorded deaths in each country, except Spain, where only comparable stratified data over an 8-week time period was available. The Global Burden of Disease database provided data on all deaths and those from pneumonia, cardiovascular disease combining ischaemic heart disease and stroke, chronic obstructive pulmonary disease, cancer, road traffic accidents and dementia.

**Findings:** Deaths related to COVID-19, while modest overall, varied considerably by age. Deaths as a percentage of all cause deaths during the time period under study ranged from <0.01% in children in Germany, Portugal and Netherlands, to as high as 41.65% for men aged over 80 years in England and Wales. The percentage of the population who died from COVID-19 was less than 0.2% in every age group under the age of 80. In each country, over the age of 80, these proportions were: England and Wales 1.27% males, 0.87% females; Italy 0.6% males, 0.38% females; Germany 0.13% males, 0.09% females; France 0.39% males, 0.2% females; Portugal 0.2% males, 0.15% females; and Netherlands 0.6% males, 0.4% females.

**Interpretation:** Mortality rates from COVID-19 remains low including when compared to other common causes of death and will likely decline further while control measures are maintained. These data may help people contextualise their risk and policy makers in decision-making.

## Background

The COVID-19 pandemic, calamitous though it is, needs to be placed in perspective. It has been eight months since the severe acute respiratory syndrome coronavirus 2 (SARS-CoV-2) outbreak was first identified^1^, and deaths globally continue to rise. As of 28 July 2020, there have been 16,523,029 cases and 654,860 directly attributable deaths worldwide. These statistics have caused widespread concern and fear^3,4^. Some of this concern is clearly justified, but some – as we have demonstrated in children – is disproportionate, given that COVID-19 caused a small fraction of deaths, even fewer than influenza^5^.

Contextualising the impact of COVID-19 in relation to other causes of death, and to mortality rates in the population, helps to gain perspective. Total mortality related to COVID-19 is the most commonly reported statistic, which has been invaluable in galvanising public health interventions^6^; however, given important differentials by age and sex, stratification of mortality data is essential^7^.

We report age- and sex-stratified mortality data related to COVID-19 and compare these with all-cause and common causes of mortality using data from the Global Burden of Disease (GBD) study^8^. We examined two perspectives: firstly, mortality from COVID-19 and other common causes of death as a fraction of all deaths, and secondly, as a fraction of the population.

## Methods

We extracted population size and COVID-19 mortality by age and sex from the National Institute for Demographic Studies website^9^ for the following countries: England and Wales, Italy, Germany, Spain, France, Portugal and Netherlands. These countries were selected due to data availability, reporting comparable age groupings stratified by sex, and comparability of location in Western Europe, with reasonably similar health care systems, economy and capacity to collect data. Available data spanned 14-16 weeks since the first recorded deaths in each country, except Spain, where only comparable stratified data over an 8-week time period was available. Furthermore, these countries have had high death rates given their average age of the population is high compared with low- and middle-income countries. These countries are therefore likely to exemplify the impact of the pandemic at the higher end of the mortality scale. Most other countries, especially with younger populations, can anticipate lower mortality.

We extracted annual age- and sex-specific death counts from the most recent Global Burden of Disease study^8^ for all causes and pneumonia, cardiovascular disease combining ischaemic heart disease and stroke (CVD), chronic obstructive pulmonary disease (COPD), cancer, road traffic accidents (RTA) and dementia; these six causes were selected as they represent common causes of death^10^.

To compare mortality estimates from the GBD with those from COVID-19, mortality rates for non-COVID-19 causes for each country were adjusted based on the number of weeks COVID-19 data were available for (Supplementary Table 1).

Data were analysed by country, age and sex with deaths related to COVID-19 and to other specific causes as a fraction of both all causes of death and population size. Data extraction and analysis was carried out by BO and checked independently by JB. Butterfly charts with stacked bars display these data graphically.

## Results

Table 1 shows mortality by cause, age and sex in the seven countries from 09/03/2020 until 09/07/2020 (for specific dates see Supplementary Table 1) and the percentage of COVID-19 deaths and other causes of death with respect to all-cause mortality. Figure 1 summarises these data.

**Table 1:**
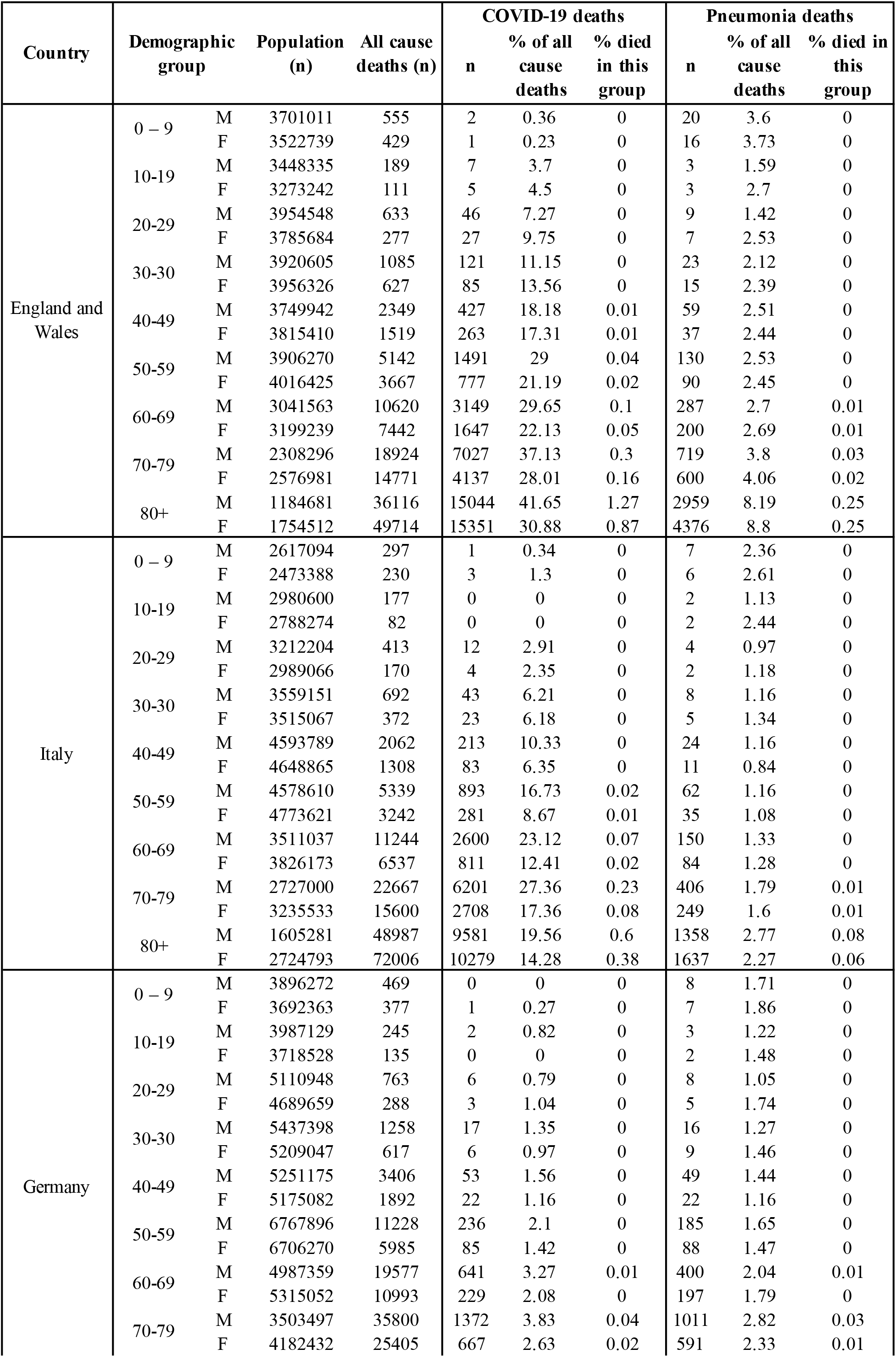
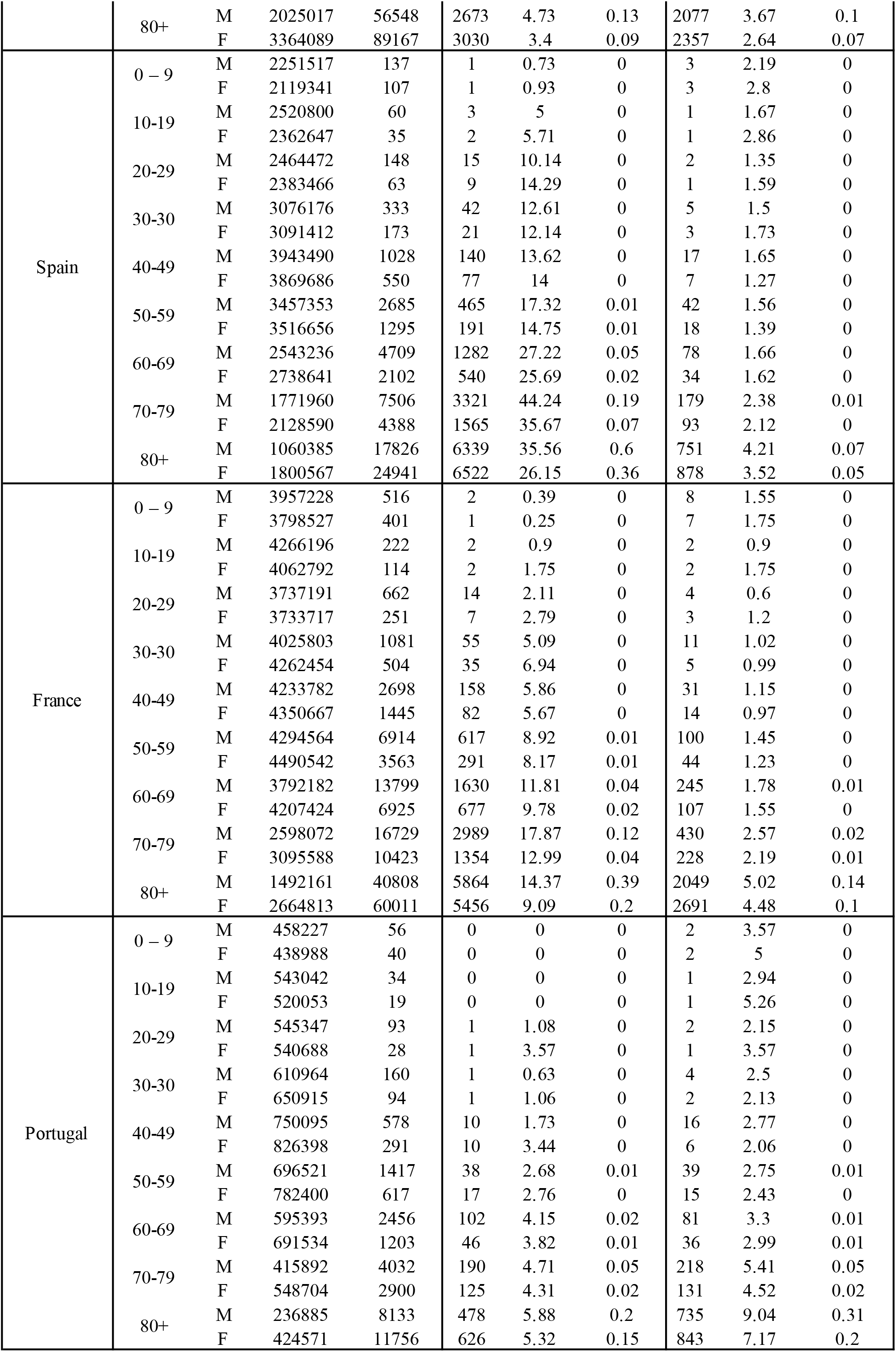
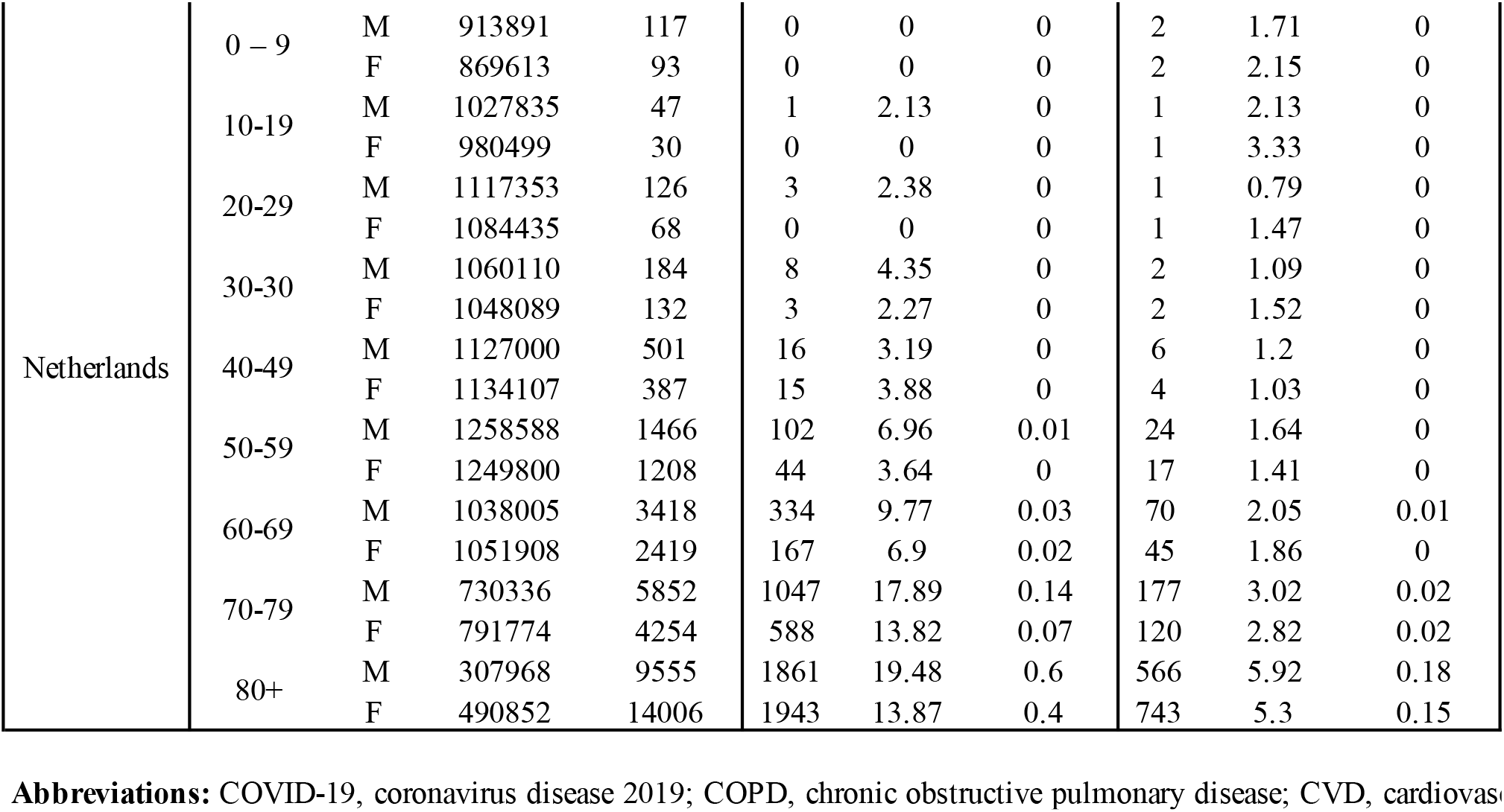
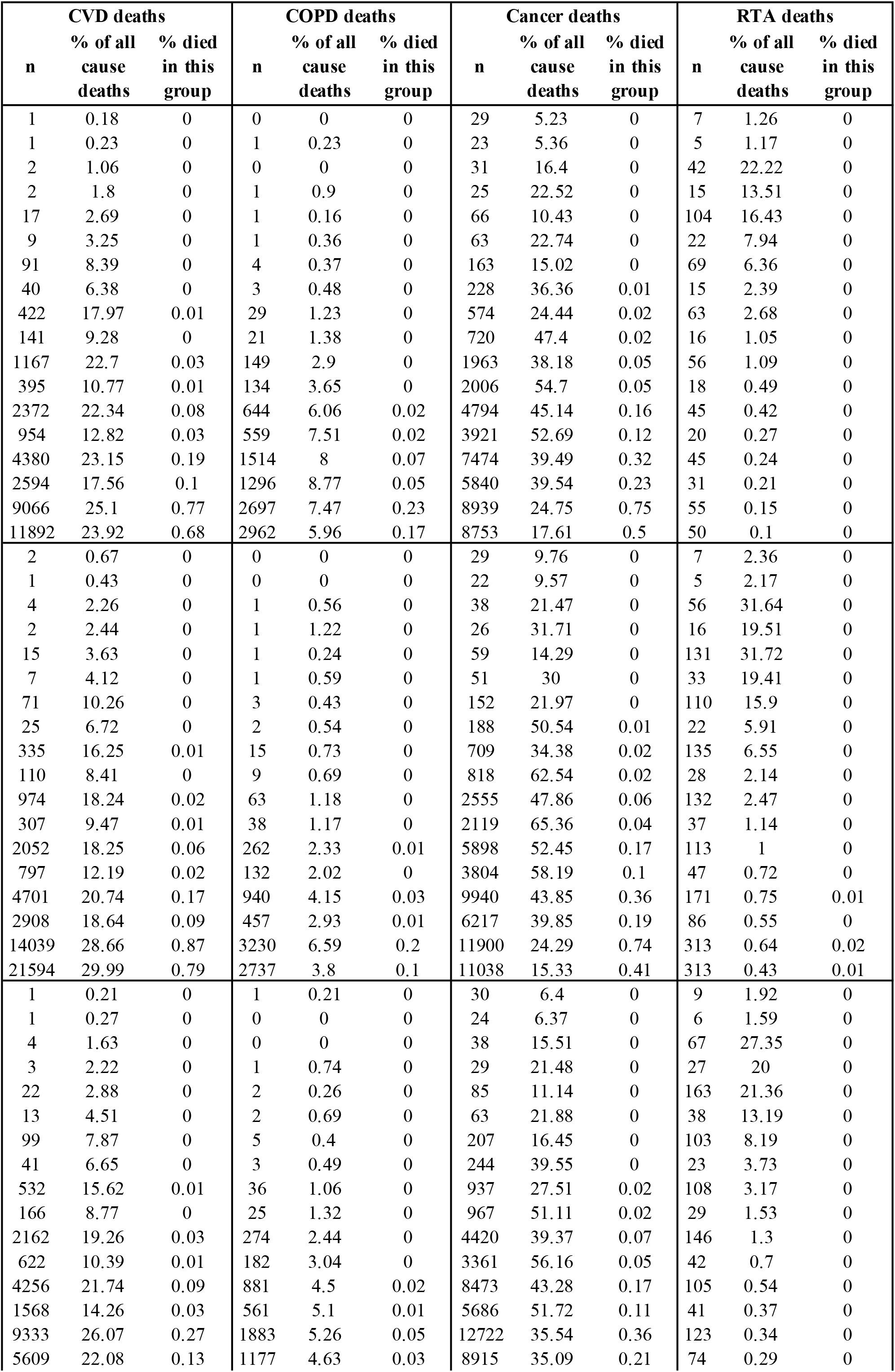
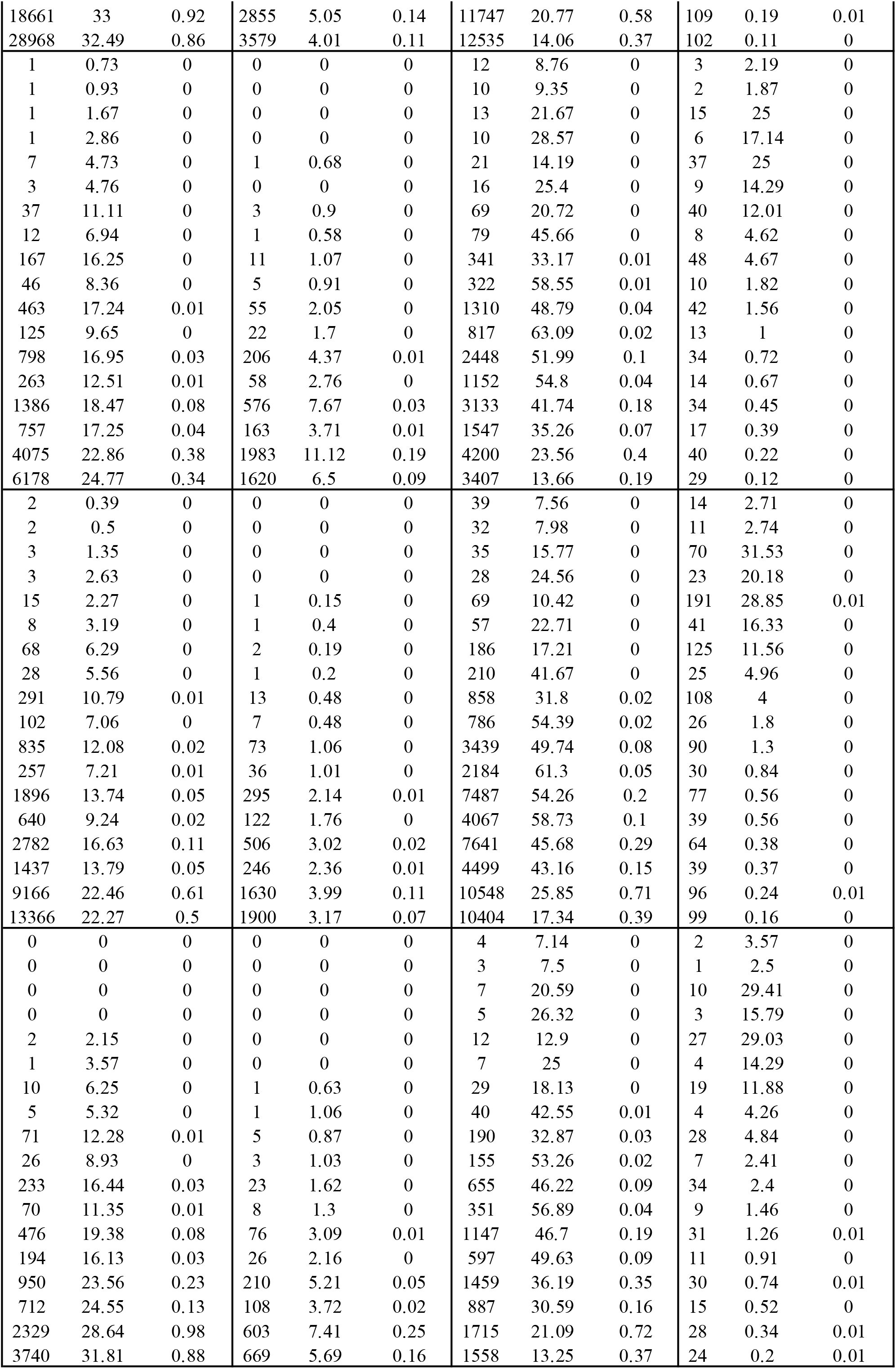
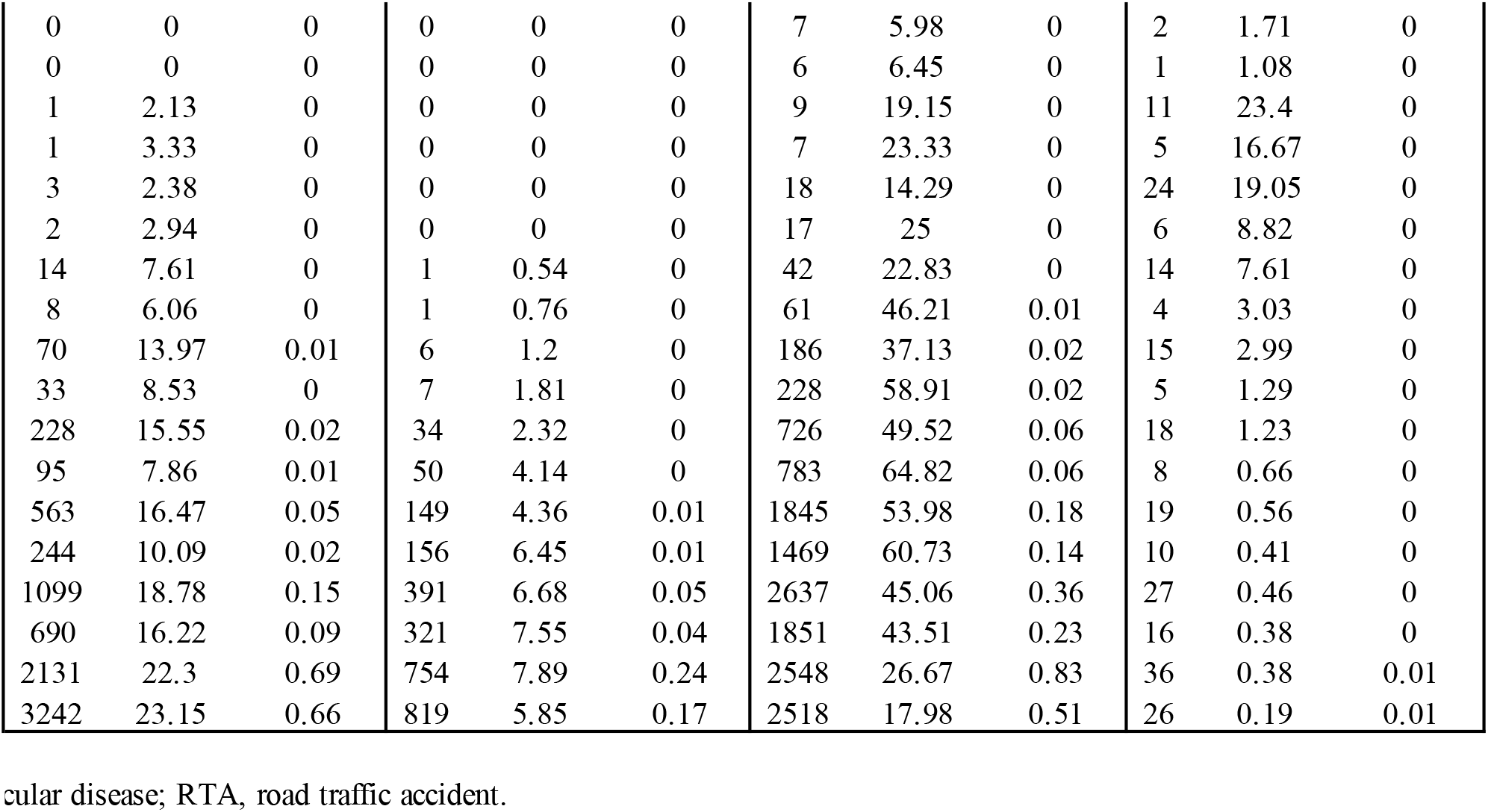
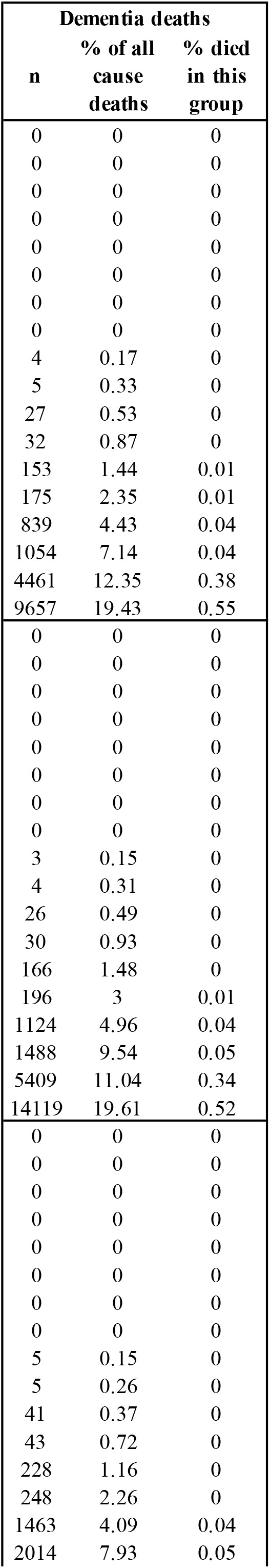
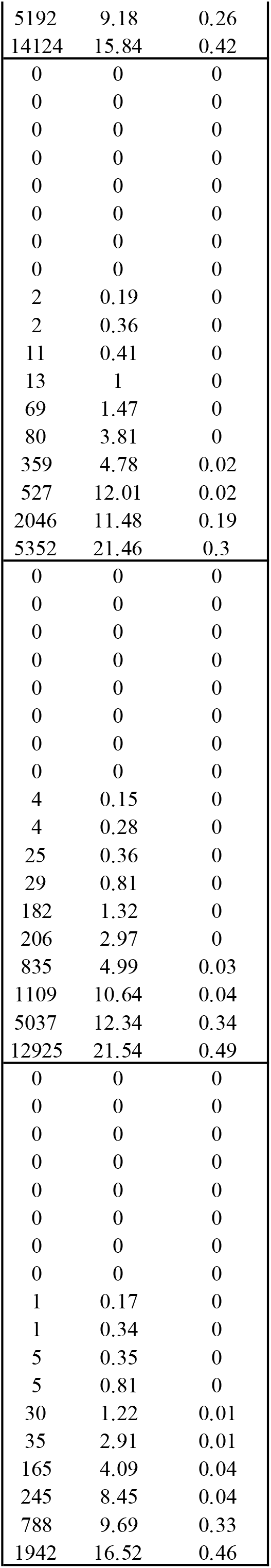
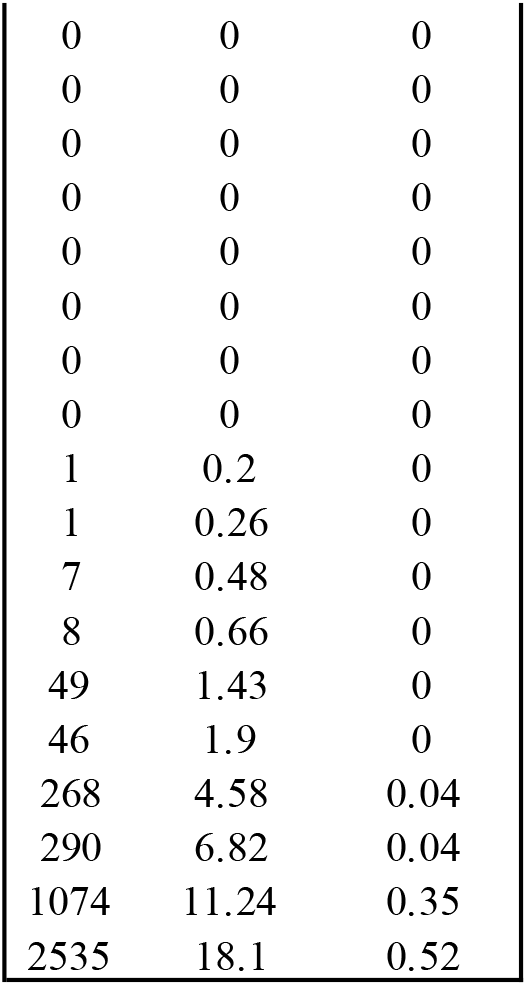
Mortality data by country, cause, age and sex: specific causes of death, including COVID-19, are shown as raw data, percentage of all-cause deaths and percentage of population for each country’s demographic group.

**Figure 1:**
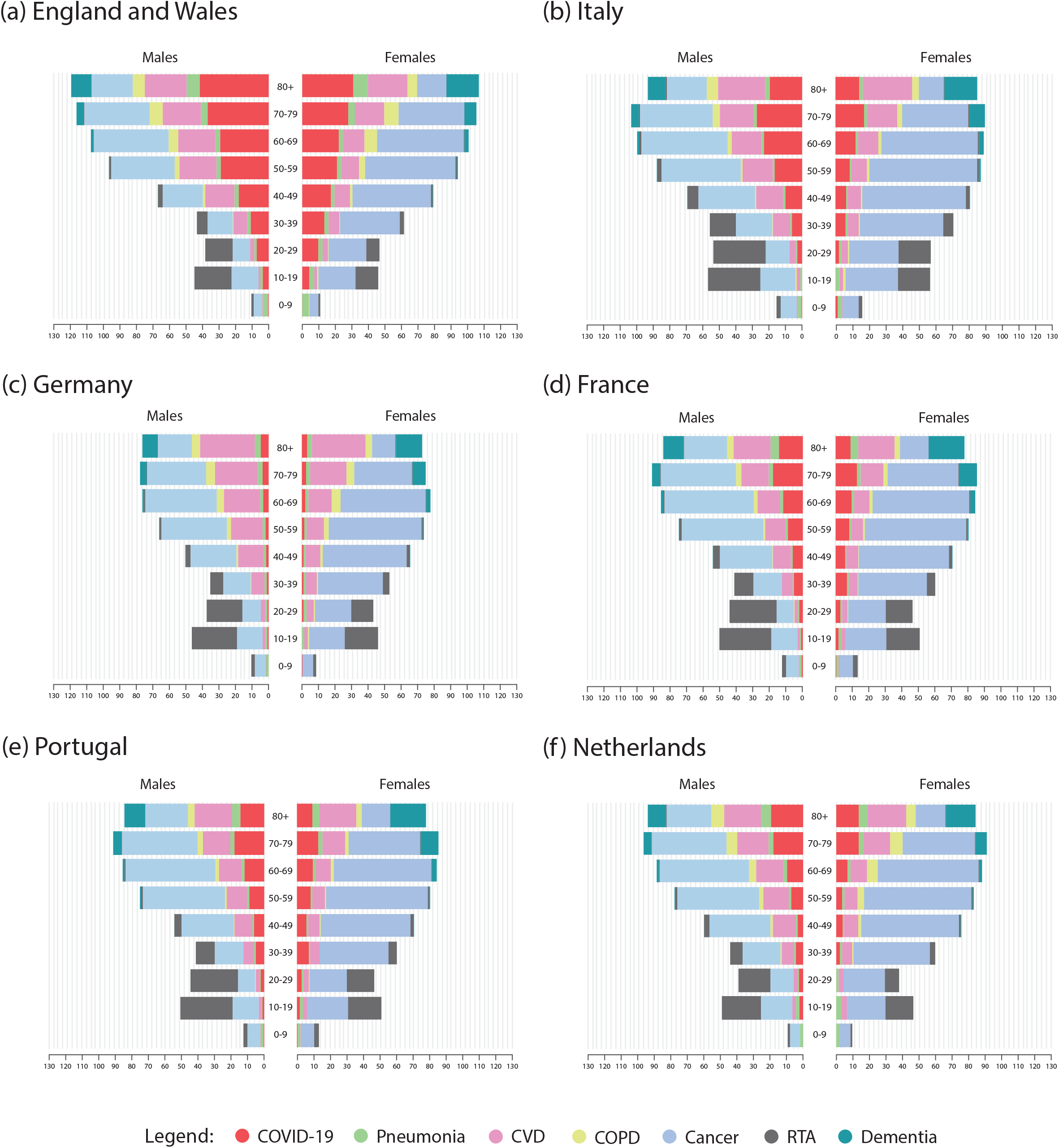
Stacked bar charts showing mortality from seven causes of death as a percentage of all-cause deaths by age and sex in six European countries.

Across all countries the number of deaths related to COVID-19 demonstrated a sharp increase with age, and there were greater numbers of deaths in males than females. Deaths related to COVID-19 represented a small proportion of all deaths overall, though this varied considerably by age being less than 0.01% in children in Germany, Portugal and Netherlands, and as high as 41.65% for men aged over 80 years in England and Wales. In groups under the age of 70, COVID-19 was never the commonest cause of death although it was an important contributor.

Figure 2 shows the percentage of the population who died from COVID-19 and the six other causes (Supplementary Figure 1 provides continuous x-axes to 100%). These figures show that, cumulatively, mortality from the six common causes of death was less than 1% in every age group, except in those aged over 80 years, where this percentage ranged from 1-4%. The percentage of the population dying from COVID-19 was less than 0.2% in every age group under the age of 80 across all countries, less than or equal to 0.1% under the age of 70 and less than 0.04% under the age of 60. In each country, over the age of 80, these proportions were: England and Wales 1.27% males, 0.87% females; Italy 0.6% males, 0.38% females; Germany 0.13% males, 0.09% females; France 0.39% males, 0.2% females; Portugal 0.2% males, 0.15% females; and Netherlands 0.6% males, 0.4% females.

**Figure 2:**
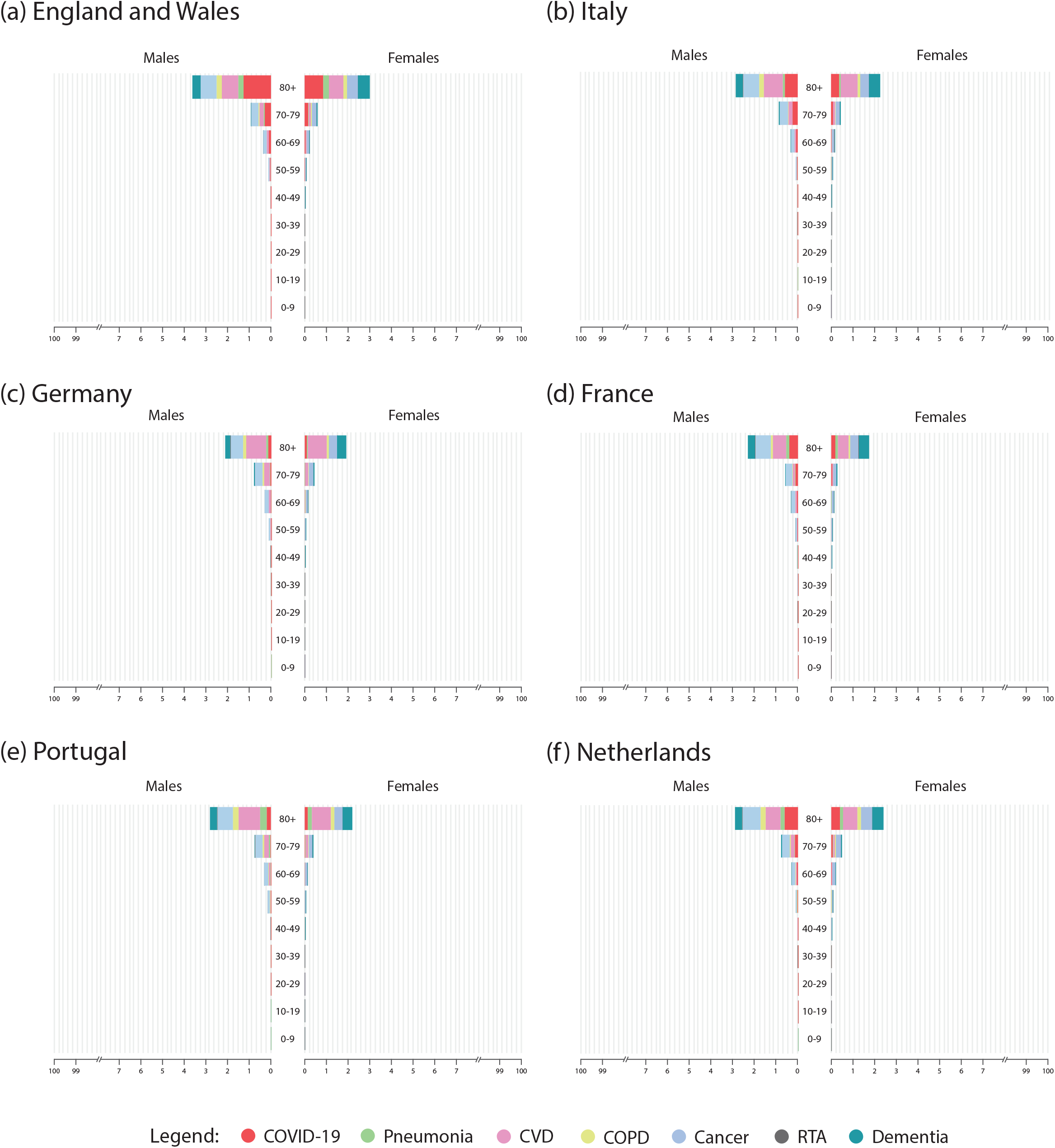
Stacked bar charts showing mortality data from seven causes of death in six countries as a percentage of the population in each demographic group. Discontinuous x-axes are used.

Graphical representation of the data from Spain are shown in Supplementary Figure 2, as these represent an 8-week time period, compared to other countries, which represent data over 14-16 weeks.

## Discussion

The COVID-19 pandemic is an international emergency warranting a comprehensive, medical, public health and economic response^11^. Our methods and analyses provide a population perspective on the pandemic in, arguably, to date some of the worst affected countries in the world. These data show that the high level of mortality is primarily seen in older adults, particularly men. However, even in the most affected groups, other causes of death were mostly commoner than COVID-19, and in all groups under the age of 70, COVID-19 did not represent the commonest cause of death. Our non-COVID-19 data are not current. Given lockdowns, with important limitations on healthcare especially for chronic conditions, it may be that mortality from these other causes will be higher in this pandemic year.

These data also highlight the very small percentages of deaths related to COVID-19 relative to population size, representing less than 0.2% in all groups under the age of 80. This mortality mainly occurred in the months of March, April and May and has largely been brought under control. We cannot forecast what future resurgences and waves of the pandemic will bring (hopefully both the incidence and mortality rates will continue to diminish) but we can see the population impact in relation to mortality so far has been modest except in those over 80 years of age. In the immediate future, the relative proportions of deaths from COVID-19 compared to other causes in these European countries are likely to decline as control measures, while being relaxed, are likely to be applied partially and intermittently for some years.

Mortality related to COVID-19 is known to be higher in males than in females and higher in older age groups and the mechanisms for these differential effects have been postulated^12,13^. Other important factors have also been recognised to lead to poorer outcomes following COVID-19 infection, including co-morbidity^14^ and ethnicity, with data suggesting that ethnic minority groups are at increased risk of death from COVID-19^15^. Though these have not been analysed in this study, ensuring a holistic approach when determining and addressing risk is important.

We acknowledge limitations of this study. We found variations between countries in proportions of deaths but have not emphasised them as data collection factors may contribute to this. For example, the COVID-19 mortality data from France represented only in-hospital deaths, whereas England and Wales also counted community deaths, including hospices and patients’ homes^9^. A further limitation is that data from Spain only represented an 8-week time span during the initial outbreak, when mortality rates were higher, as their data reporting methods changed beyond May^9^, hindering access to comparable data since then. Defining COVID-19 mortality rates is also contentious, as data pertains to clinically apparent PCR-positive infections, underestimating true mortality^16^. Furthermore, there may be several reasons why the mortality totals exceed 100 in England and Wales, Italy and Spain. The cause of death reporting may include more than one of the listed causes of death in this study, therefore leading to an overestimation of the cumulative totals. Without access to real-time mortality data on all causes, we are also unable to assess the ongoing effect of the pandemic on mortality related to other causes, such as cancer and cardiovascular disease, which may rise as healthcare resources have been both curtailed and diverted^17^.

Our data provide an important public message for policymakers, healthcare workers and the public, who are trying to understand the impact of COVID-19 and the risk of dying. Misinformation has been a problem, perpetuating public fear and anxiety, impacting on the increasing burden of adverse mental health during the pandemic and even contributing to suicide risk^18,19^.

By presenting and interpreting population perspectives on mortality related to COVID-19 compared with other common causes of death, stratified by age and sex, we have provided perspectives to allow policymakers, professionals and the media to tailor both communications and interventions to manage the pandemic, including the level of anxiety and fear provoked by previously published mortality statistics, primarily daily and cumulative totals. Similar analyses are required globally. New work is required to incorporate morbidity to produce a broader perspective on the true health impact of COVID-19.

## Data Availability

Data sharing not applicable.

---

**Supplementary Figure 1:**
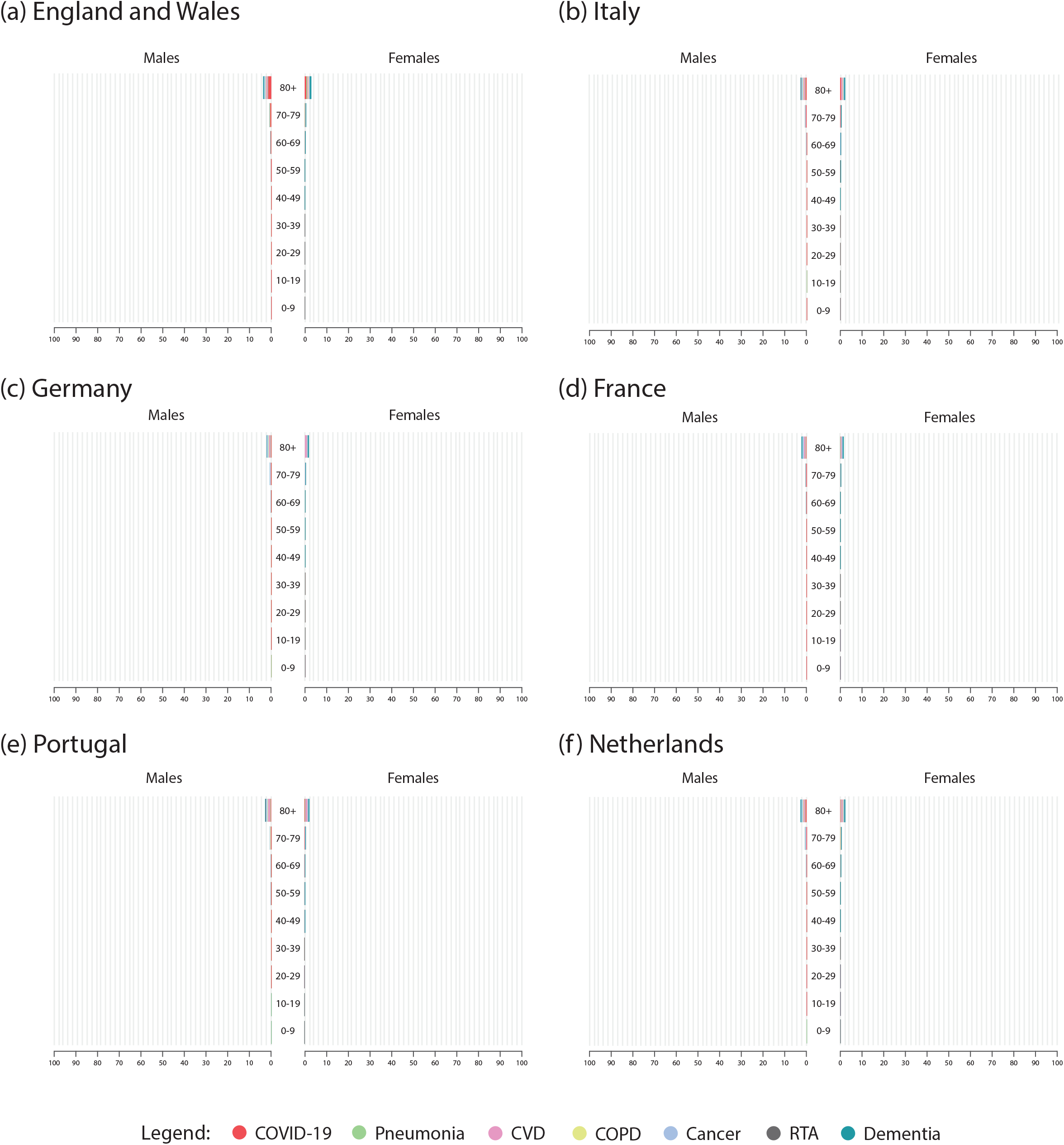
Stacked bar charts showing mortality data from seven causes of death in six countries as a percentage of the population in each demographic group. Continuous x-axes are used.

**Supplementary Figure 2:**
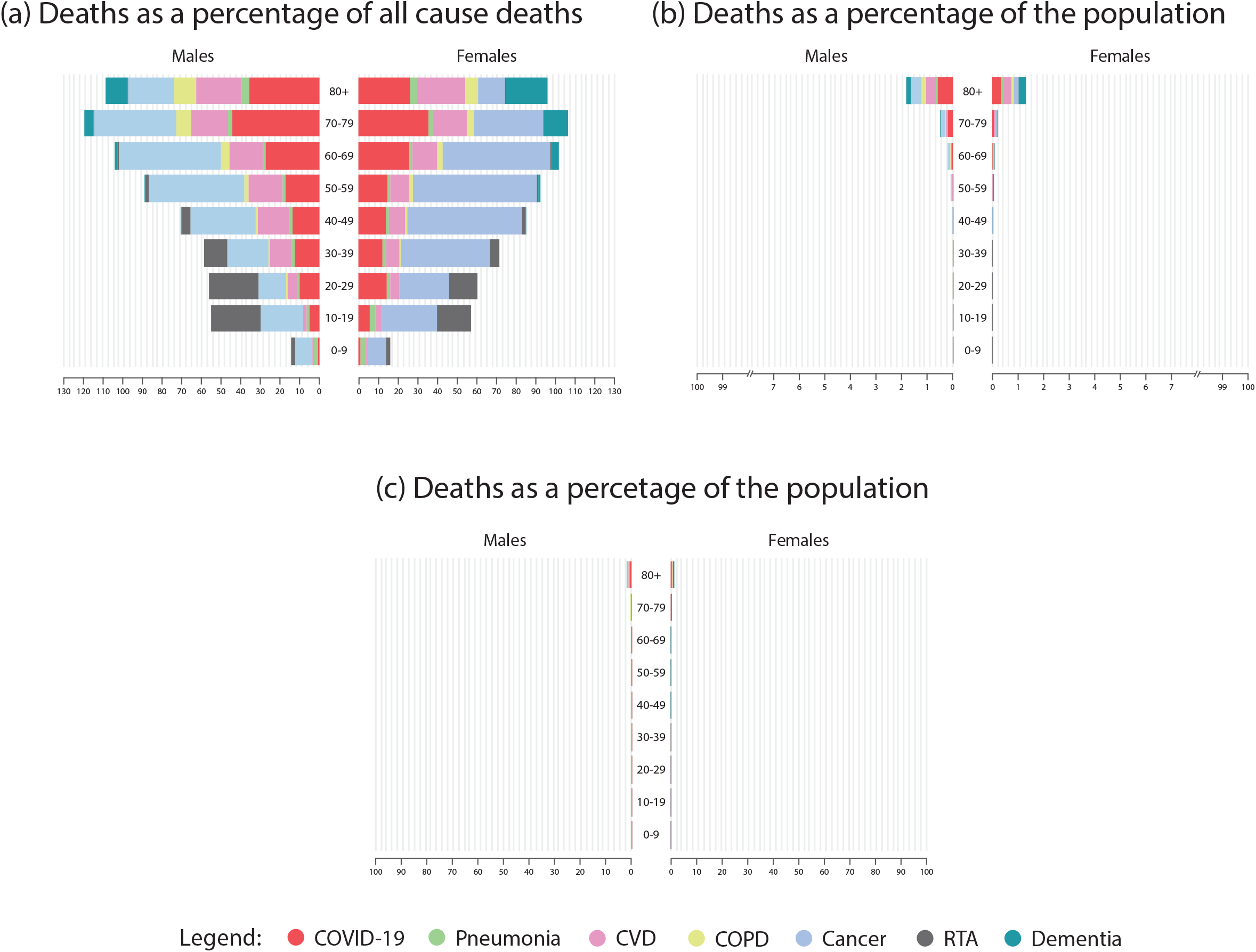
Stacked bar charts showing mortality data from Spain (a) as a percentage of all cause deaths, and (b) and (c) as a percentage of the population.

**Supplementary Table 1:**
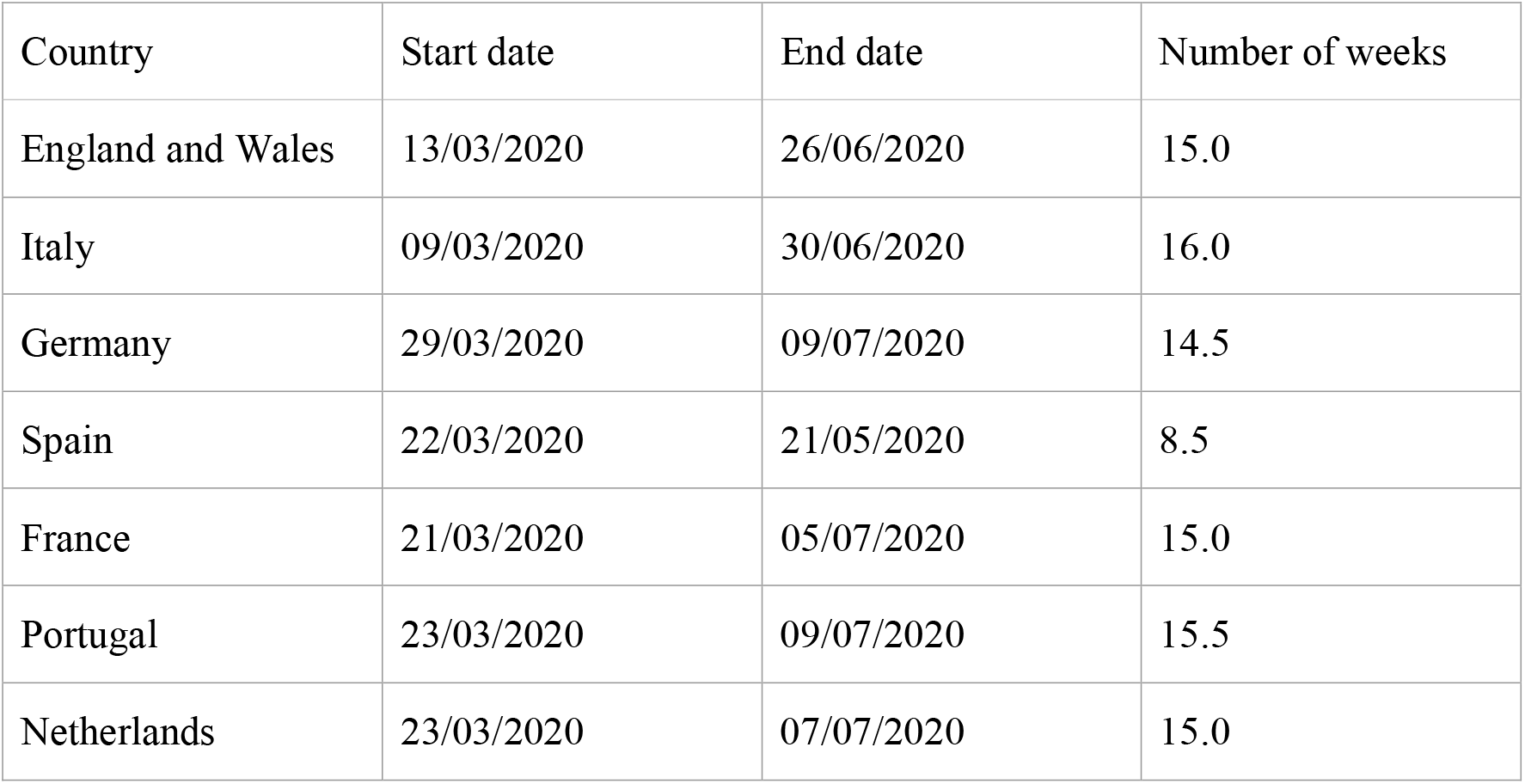
The time period for which age- and sex-specific COVID-19 mortality data was available for each country and the number of weeks this spanned.

## Contributions

RB conceived the study. SB and JB developed the methodology, which was expanded by BO. BO carried out data extraction, which was checked independently by JB. BO carried out the data analysis. All authors contributed to the interpretation of the data. BO wrote the first draft of the manuscript, which was substantially edited by all authors. All authors approved the final version. All authors had access to the data and are responsible for data integrity and completeness.

